# Harnessing digital tools for the management of large-scale larviciding operations: a pilot of the ZzappMalaria software system in Obuasi, Ghana

**DOI:** 10.1101/2024.03.07.24303784

**Authors:** Ignatius Williams, Kwame Desewu, Samuel Asiedu, Nicholas Ato Egyir, Dan Gluck, Arnon Houri-Yafin, Yonata Fialkoff, Arbel Vigodny

## Abstract

Larval source management (LSM) has played a critical role in historical malaria elimination endeavors, yet its application in tropical settings has been hampered by operational hurdles and limited effectiveness. To address these issues, ZzappMalaria Ltd. developed a digital platform for the planning and implementation of LSM campaigns, the features and field efficacy of which are discussed in this paper.

Piloted in collaboration with AngloGold Ashanti Malaria Control Ltd. (AGAMal) in Obuasi, Ghana, the system, comprising a planning tool, a mobile app for fieldworkers and an online dashboard for monitoring and worker management, facilitated the detection of more than 4,000 water bodies and reduced the mosquito population by 62% for a cost of $0.24 per person protected (PPP). The data available does not provide sufficient evidence to conclusively determine whether a reduction in malaria cases occurred; nevertheless, the results underscore the potential of digitization to surmount the operational challenges of large-scale LSM campaigns.

## Background

Larval source management (LSM) is an effective tool for reducing *Anopheles* mosquito populations, targeting both indoor and outdoor-biting species.^[1]^ LSM was the mainstay of several successful malaria elimination operations around the world in the 1930s and 1940s.^[2]^ However, because a single water body can support a large number of mosquito larvae, the success of LSM depends greatly on the proportion of water bodies that can be detected and treated. The operational difficulties associated with LSM have often led to poor intervention coverage and an insufficient impact on the mosquito populations, preventing LSM from serving as a standalone.^[3]^ As a result, the current WHO guidelines recommend LSM as a supplementary intervention to long lasting insecticidal nets (LLIN) and indoor residual spraying (IRS), and only in locations where mosquito breeding sites are “few, fixed and findable,”^[4]^ i.e., where there is a limited number of water bodies, which are permanent and can be easily detected.

The effectiveness of IRS and LLIN, however, has been hindered by various factors, including the emerging resistance of mosquitoes to insecticides^[5]^ and an increased tendency for outdoor biting.^[6]^ As a result, there is a growing need for integrated vector management (IVM), i.e. the combination of several methods and tools in the same location. ZzappMalaria has developed a digital system, comprising a mobile application and an online dashboard, with the dual goal of overcoming the operational challenges associated with larviciding, as well as optimizing its integration with other intervention methods.

ZzappMalaria’s software facilitates the management of large-scale LSM interventions, ensuring detection of a high proportion of the potential mosquito breeding sites, as well as their regular treatment with larvicides or other methods.^[7]^ LSM operations, if executed and monitored properly, have the potential of being more cost-effective than IRS and LLIN, and may serve as a significant supplementary method for each.^[8]^ This is especially true in light of the growing rate of urbanization in sub-Saharan Africa, since the cost of larviciding per person protected is inversely related to population density. LSM is also relevant for areas with longstanding LLIN or IRS operations, where it can offer a way to boost stagnating declines in malaria prevalence, and help mitigate the impact of insecticide resistance in mosquito populations.^[9]^

This paper presents the functions of the Zzapp system and reviews a pilot conducted in collaboration with AGAMal in Obuasi, Ghana to test its usability for managing large-scale larviciding operations. It examines the system’s components, functionalities, field use, and cost-effectiveness in reducing mosquito populations and malaria incidence. The paper concludes by discussing the potential benefits of digital LSM in operational, managerial, and scientific contexts, as well as the ways in which it can promote IVM operations.

### Overview of the Zzapp technology

Zzapp is a software system that facilitates the planning, management and monitoring of malaria vector control interventions. In the context of LSM, the system was designed to increase the coverage of water body mapping and ensure their regular treatment against mosquito larvae . The system comprises a web-based dashboard where operations are planned and monitored by administrative staff, and a map-based mobile app that guides workers in the implementation of interventions. The mobile application operates on Android smartphones, and requires a GPS sensor, a compass, and a camera. Internet connectivity is required initially to download the maps, and periodically to sync data and receive assignments; however, the app is designed for offline use in the field.

LSM operations may be divided into two phases, mapping and treatment. In the mapping phase, the system first outlines an initial area of treatment, determined by the inhabited area of the targeted community. The system applies machine vision analysis to satellite imagery, automatically identifying any built structures and grouping them into clusters with clustering algorithms. To accommodate the flight range of mosquitoes, an additional ‘buffer zone’ around the inhabited area is added. The size of the buffer zone is modified to include areas with a high probability for the presence of water bodies based on topographical and satellite imagery analysis. After finalizing the intervention area, the system subdivides it into operational units, known as ‘villages,’ typically measuring half a square kilometer each. Team leaders in the field use a mobile app to further divide these villages into sectors, with each assigned to a specific fieldworker. The app helps delineate sector boundaries, aiding fieldworkers in navigating their allocated tasks.

The progress of fieldworkers in mapping activities is recorded on the app by a GPS-based terrain coverage feature, visualized on the map as a yellow overlay. This overlay consists of semi-transparent squares, corresponding to 10 square meters each, that cover areas that the field workers have visited (figure 1). Depending on the level of accuracy required for the project, the radius around the worker’s GPS location that is marked as surveyed for each GPS point can be modified, resulting in very thorough mapping when the radius is small (since the worker has to visit every 10 square meter pixel to turn it yellow), or less thorough when the radius is larger. This mapping radius reflects the distance from which field workers are expected to detect a potential breeding site.

**Figure 1:**
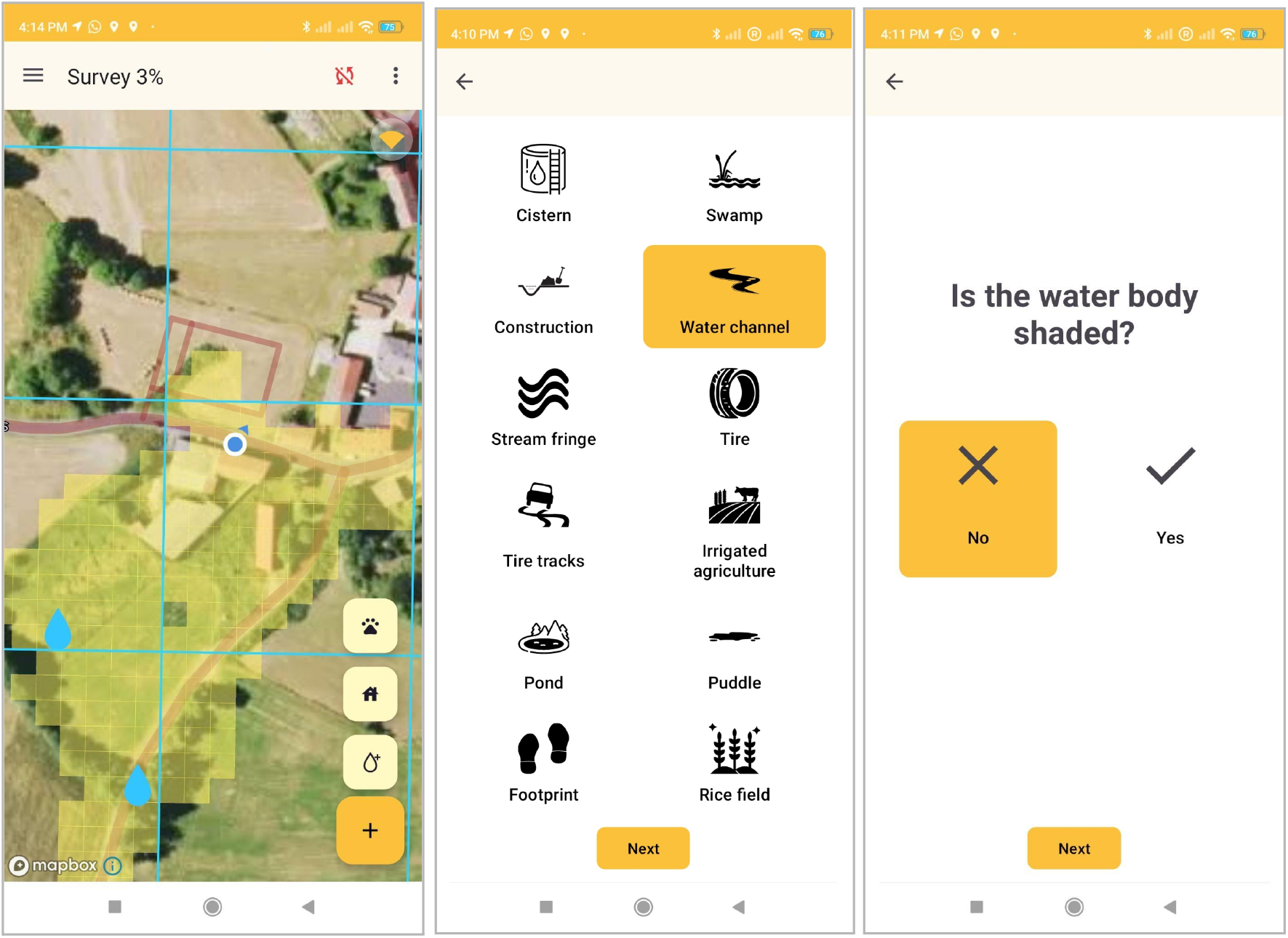
Screenshots from the Zzapp mobile application showing the mapping coverage feature visualized as semi-transparent yellow pixels covering areas visited by the field worker, and two sample questions from the water body questionnaire used by field workers to report water bodies they encounter in the mapping of potential breeding sites.

The mapping radius, and hence the thoroughness and cost of breeding site mapping, are set by operation managers before field operations begin, based on the availability of resources and personnel, required thoroughness, duration of the mapping phase and other operational considerations. The terrain coverage tracker aids in monitoring of field workers and ensuring complete coverage of the intervention area. In this study, the mapping radius varied depending on the topography of the area being mapped. For terrain identified as suitable for water bodies, such as valleys^[10]^, a narrow 10-meter mapping radius was selected to ensure thorough mapping. For all other regions, a wider 17-meter mapping radius was adopted to optimize resource usage.

The app also features a ‘roam out’ alert, which notifies fieldworkers when they have exited their designated treatment zone through a vibration notification on their mobile device. This is to avoid duplicate mapping and increase fieldworks’ individual responsibility. When fieldworkers locate a water body, they create a report on the app that includes a photo, GPS location and additional details captured through a questionnaire (figure 1). These data appear on the dashboard as icons on a hybrid satellite and street map (figure 2). The system also includes a module that enables the integration of water body location data obtained by drones, which was not used in the study reported here.

**Figure 2:**
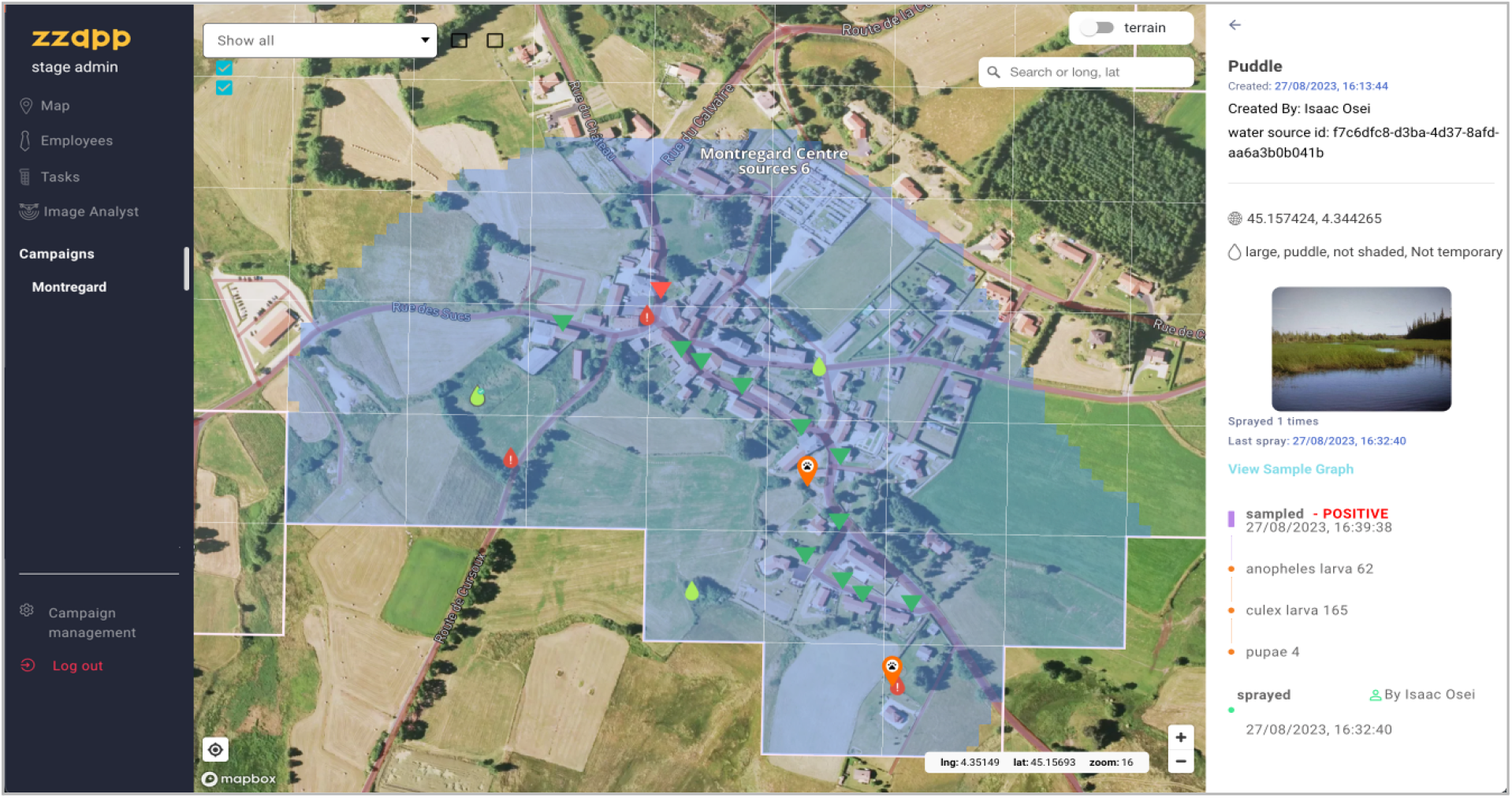
A screenshot from the web dashboard showing a sample area following mapping and treatment tasks. The mapping coverage on the dashboard is visualized in blue (compared to the app, where it is yellow). The droplet icons indicate water bodies - green water bodies are ones reported as treated (within the treatment expiration period), and red water bodies indicate issues preventing treatment. Other icons shown indicate private property visited by field workers (triangles), and animal shelters (paw icons); however, these elements were not used in the pilot discussed herein.

During the treatment phase, water bodies that were located in the mapping phase are regularly treated with larvicide (or by draining/covering the water body). The frequency of treatment depends on the duration of larvicidal activity of the material used once it is applied onto the breeding site. The expiration period is influenced by the type and concentration of the larvicide material used, as well as weather conditions (e.g., rainfall that may wash away the larvice from water bodies).^[11]^ Assignments during this phase are automated via a K-means based algorithm, which distributes the tasks based on the proximity of water bodies to each other and to the fieldworkers, while also ensuring an even workload among the team members. After a water body has been treated, its icon on the system’s interface changes from blue to green. The icon reverts back to blue after an expiration period, set to 14 days for the operation discussed herein. Field workers can also report issues, signifying that they were unable to treat a specific water body (e.g., because it dried up or because access to it was denied by the property owner).

After the completion of a mapping or treatment assignment, team leaders review the work performed. They can either approve the assignments as complete, or send the workers back to the field if additional work is required. This iterative process ensures high coverage and effective treatment of identified water bodies. The system also supports entomological sampling of the water bodies that are reported during the mapping phase. Sampling is done prior to the treatment phase to create a baseline estimation of larval densities, and during the treatment phase for quality assurance of the larvicide application process and in order to assess the overall progress of the operation. Sampling results are recorded on the app, including information about the number of larvae and pupae that were found, their genus, and their developmental stage. This information is integrated with other information about the breeding sites that is uploaded during the mapping stage. In addition to quality assurance, the Zzapp system can simulate the larval positivity data to create a spatial model of malaria transmission in a given village/town. To build the model, the system integrates data on the location and larval positivity of water bodies, with locations of houses, extracted from satellite imagery using machine vision or retrieved from databases such as OpenStreetMaps or the Open Building project.

In addition to facilitating the execution of operations, the system also supports their monitoring. The dashboard displays mapping coverage data allowing managers to spot gaps in the ground survey and evaluate their significance. In addition to the map, the dashboard produces daily reports that summarize the activities by users and teams. These tables can be used to easily identify underperforming workers and other issues. Finally, the tables, and the dataset of the system as a whole, can also be used as a research tool to gain insight on mosquito dynamics and generate recommendations about management of LSM and IVM operations.

## Methods

### Study site

The operation took place in the Ashanti region of Ghana, covering a 68.3 km^2^ area that included the entire mining town of Obuasi and several adjacent communities (figures 3 and 5). This area is home to approximately 208,000 residents.^[12][13]^ The reported malaria incidence in Obuasi for 2019 was 97.1 cases per 1000 persons (20,201 confirmed cases). The dominant *Anopheles* species in the area is *gambiae* with a 50% indoor-biting rate (AGAMal, unpublished data, 2019), followed by *funestus* with an unknown indoor-biting rate. In addition to the typical *Anopheles* breeding sites (swamps, puddles and artificial water bodies), the municipality is also surrounded by “galamsey”, namely holes dug by illegal miners that may fill up with water and create breeding sites for mosquitoes. The Ashanti region has a tropical climate, and rain can be expected throughout the year. However, the area usually experiences two drier periods each year: the short dry season typically occurs between October and November, and the long dry season takes place between December and February.

**Figure 3:**
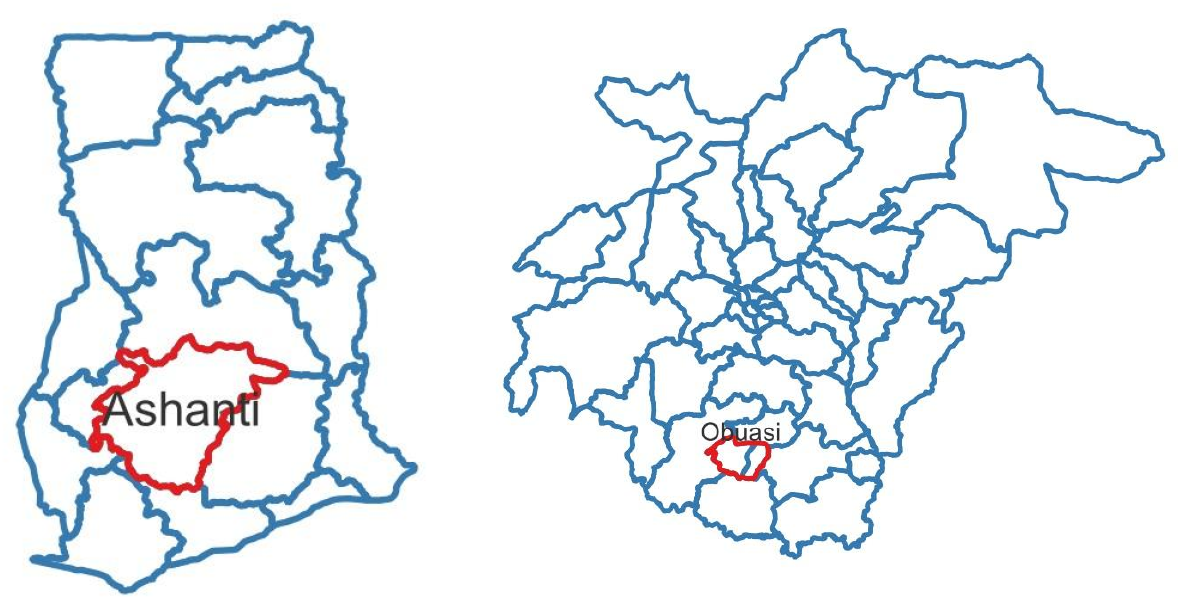
The pilot took place in Obuasi, the second largest city in the Ashanti region of Ghana.

**Figure 4:**
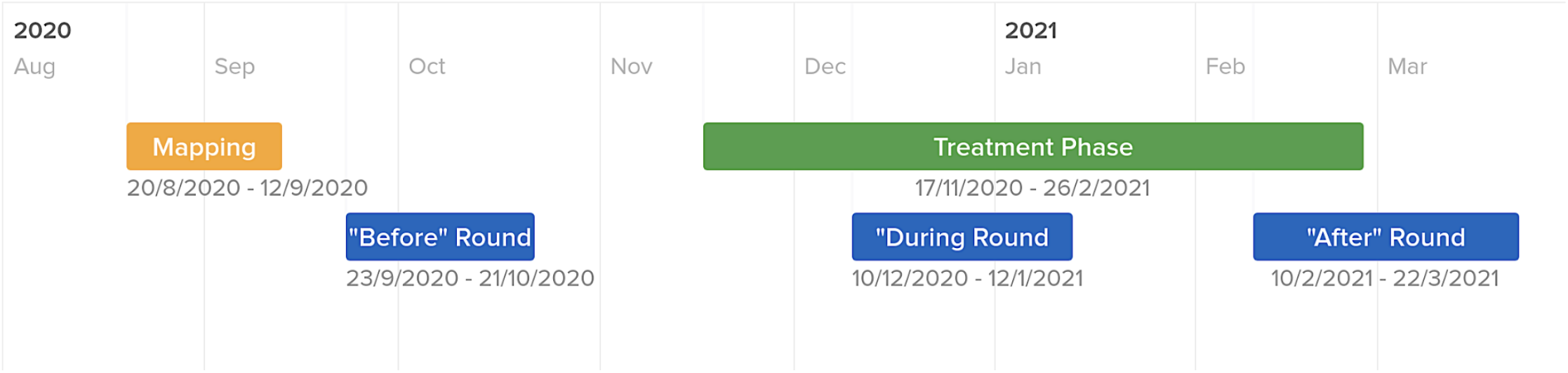
Operation timeline

**Figure 5:**
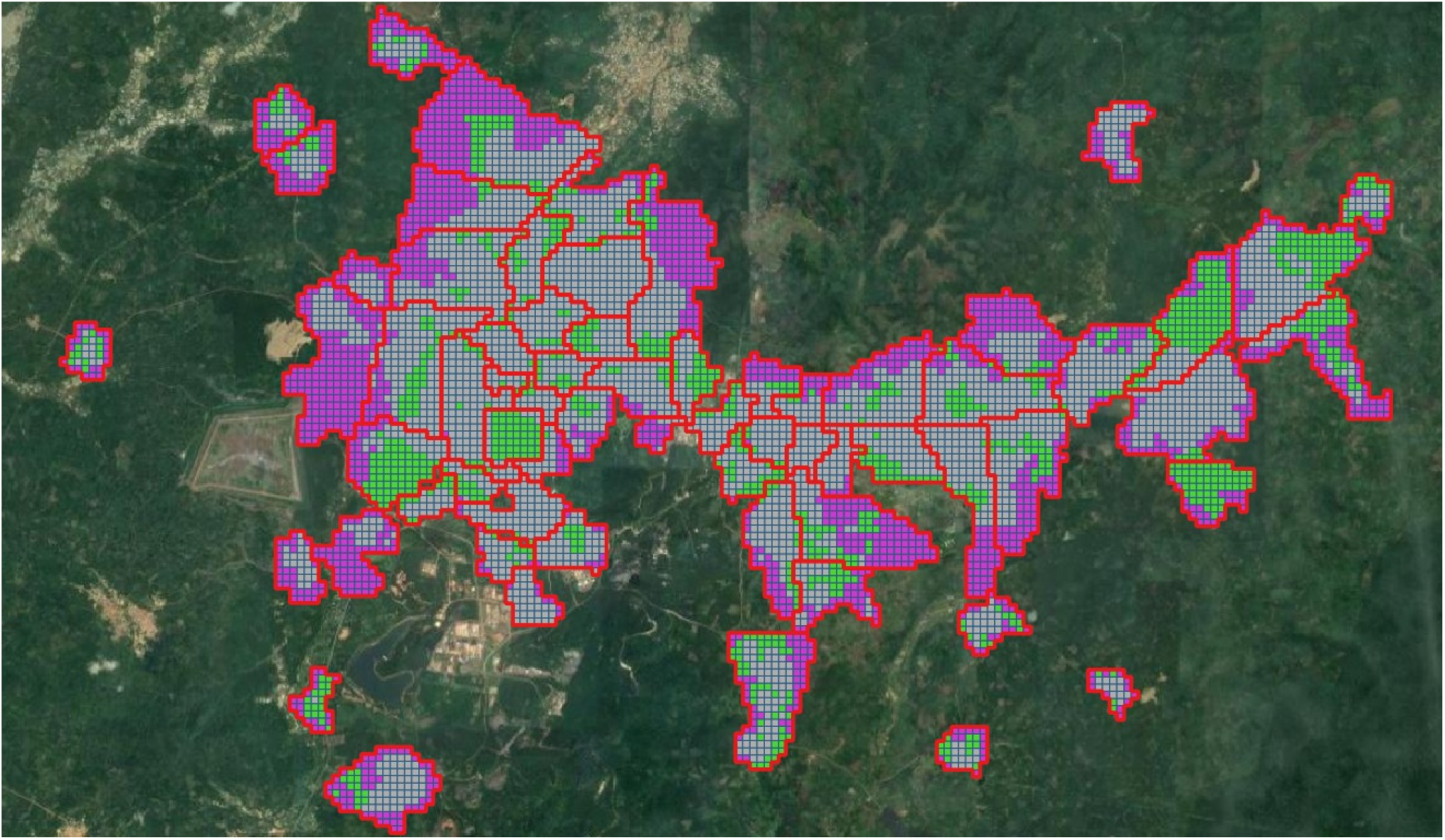
Map of the designated intervention area, color-coded by levels of urbanization. Gray: densely built areas, 83% mapping coverage. Green: areas within the city with low structure density, 50% mapping coverage. Purple: areas around the city that are uninhabited but were assigned for mapping, 18% mapping coverage.

**Figure 6:**
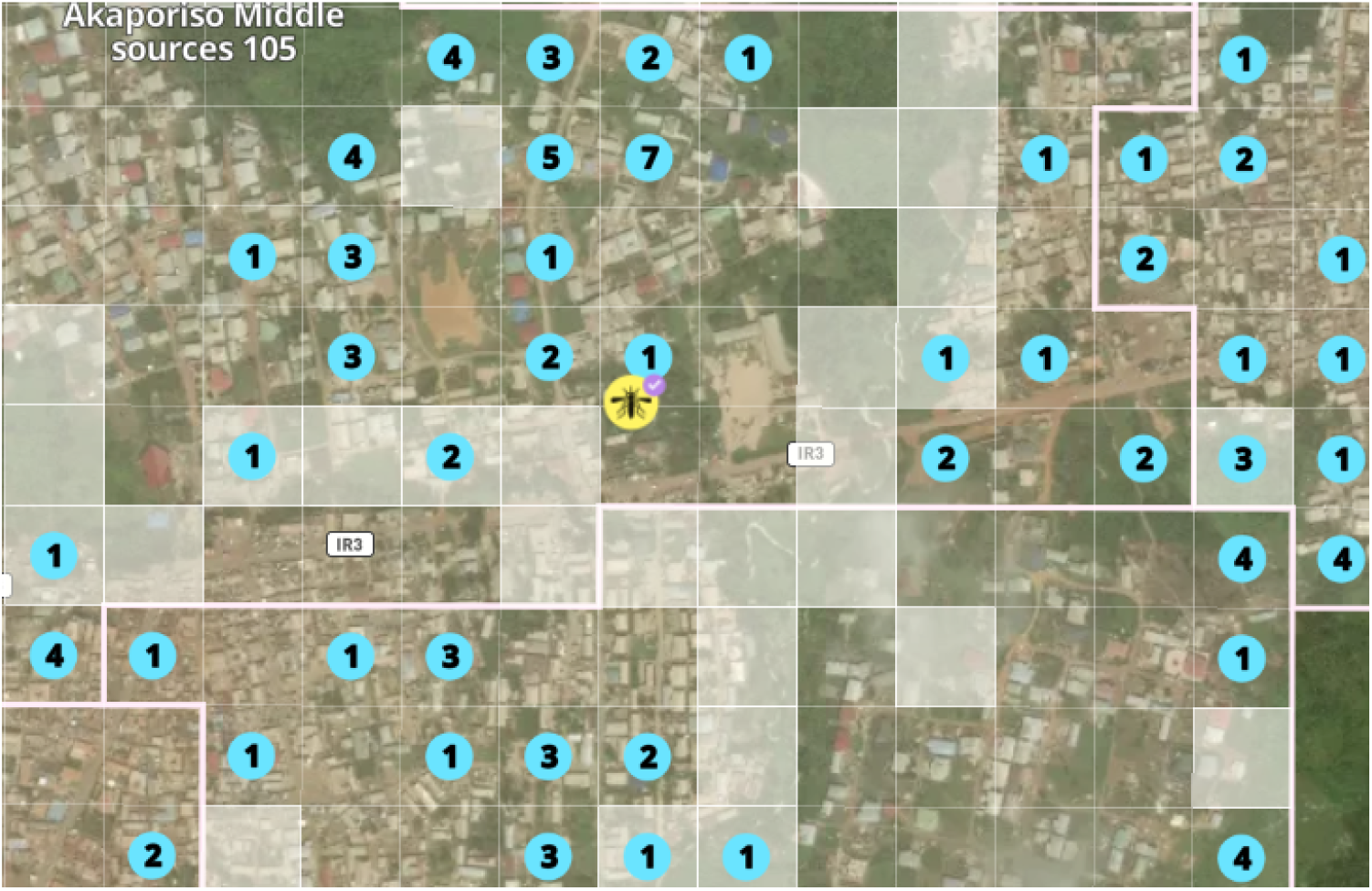
For each light trap (yellow mosquito icon), the system can calculate its distance from surrounding water bodies, their type, positivity (if sampled), and the exact dates when they were treated.

### Study Design

#### Mapping coverage trial (2018)

In 2018, ZzappMalaria and AGAMal conducted a short randomized controlled trial to test the system’s usability and effectiveness in facilitating the detection of water bodies. This small trial did not include treatment of water bodies.

During the trial, ten fieldworkers were randomly divided into two equal teams. Their goal was to detect water bodies within an area measuring 1,200m x 600m. The trial was carried out over two days, with team 1 tasked with searching area A on the first day and area B on the second day. Conversely, team 2 started with area B on day 1 and switched to area A on day 2. Throughout the trial, one team used the app to aid in water body detection, while the other team employed traditional methods without app assistance. Data for both groups was extracted from the mobile app database for the app arm, and from a combination of paper forms and a GPS logging app for the traditional mapping arm.

#### Larviciding intervention (2020-21)

In order to test the practical usage of the system, ZzappMalaria collaborated with AGAMal in 2020 to conduct a digitized LSM operation. The goals of the operation were threefold: (1) reducing malaria cases; (2) reducing the mosquito population; (3) examining the system’s contribution to management of LSM operations and comparing the operation’s cost to that of a similar-scale IRS operation.

#### Intervention Design

Before the start of the operation, fieldworkers and managers received training, which, due to COVID-19 restrictions, was conducted remotely. The training focused on the use of the mobile app and dashboard, and stressed the importance of high coverage of water bodies for the success of LSM operations. Aside from the Zzapp system (the software as well as the smartphones and computers), the equipment (e.g., sprayers) was standard and approved for LSM operations. The larvicide used was a Bti-based product (VectoBac^®^ WG).

During the mapping phase, 30 fieldworkers surveyed the intervention area over the course of 17 days, between 20/08/2020 and 17/09/2020. Water bodies were detected and reported on the Zzapp mobile app, using Infinix Hot 10 mobile devices running an Android operating system. In the treatment phase, lasting between 17/11/2020 and 26/02/2021, the water bodies were treated with Bti by 15 fieldworkers, with the goal of completing each treatment round within 2 weeks. Prior to treatment, fieldworkers visually inspected the water bodies for the presence of larvae. If fieldworkers suspected that larvae were present, the entomology team would be called to sample the water body prior to treatment.

#### Data collection and analysis

To calculate the impact on malaria cases, we relied on official data from the ministry of health. To calculate the impact on *Anopheles* mosquito population, we estimated the reduction in mosquito densities based on collections from indoor mosquito light traps (CDC light traps, J. W. Hock Company). Mosquito population density was assessed before any water bodies were treated (23/09/20 - 21/10/20) and towards the end and shortly after the treatment phase (26/02/2021 - 22/03/2021). A total of 24 collection sites were monitored, with 12 in the intervention area and 12 in a control community. The selection of locations was based on the availability of volunteer households. Each site was sampled for 15 nights per round, resulting in a total of 720 collections. The analysis involved calculating the before-and-after ratio for each trap and comparing the average change in mosquito counts between the intervention and control areas.

Mosquito densities were calculated by summing the mosquito count from the 15 nights in each collection round for each site. The change per site was determined by dividing the overall count in the collection round after the intervention by the overall count in the collection round before the intervention. The overall change for each arm (intervention/control) was calculated as the median change across all 12 collection sites in each arm. Traps with less than 5 mosquito collections in the “before” period were excluded, to avoid division by small numbers.

Median values were used as the primary measure to ensure robustness. Mean values are reported as well. Since the values were not expected to follow a normal distribution, bootstrapping was employed to calculate the confidence interval. This involved excluding one collection site in each arm at a time and recalculating the median ratios, repeating the process for all possible pairs of collection sites. The 95th percentile of the resulting median ratios was reported as the 95% confidence interval.

The overall financial cost of the operation was calculated using the ingredients approach, by identifying each activity involved and adding their costs (similar to Worrall^[14]^). Direct expenditures, such as larvicide material and field labor, were extracted from AGAMal financial records. Costs not readily available as expenditures were estimated as appropriate. Costs were converted to USD based on the exchange rate on 1/1/2020.

## Findings

### Mapping coverage trial (2018)

The results of the mapping coverage trial showed improved mapping in the team utilizing the app. The app not only increased the count of valid water bodies by 28% (82 vs. 64), but it also significantly decreased the instances of invalid water body reports— including duplicate entries, incorrect reports, and reports of water bodies situated outside the sector boundaries. This combination of increased detection of valid water bodies and reduced false reporting is crucial, since it has an impact on both increasing the mapping coverage obtained, as well as reducing the resources required to achieve that coverage.

### Larviciding intervention (2020-21)

Throughout the operation, both managers and fieldworkers expressed satisfaction with the system, highlighting its user-friendly nature, the enhanced detection and treatment of water bodies, and the ease of data entry, storage, and organization compared to manual reports. Valuable feedback was provided by fieldworkers and managers, who also shared ideas on improving the detection and treatment of water bodies. For instance, it was discovered that a feature in the app allowing team leaders to allocate assignments to fieldworkers via Bluetooth without internet connectivity, encountered occasional connection issues. As a result, this feature was replaced by a less flexible but more reliable alternative of assignment allocation directly from the dashboard.

The mapping coverage for different parts of the intervention area was calculated from data collected by the coverage tracker feature of the app. The image below (figure 5) is a map of the designated intervention area, color-coded by level of population density, with each type analyzed to determine mapping coverage. As seen in the map, coverage in the densely built area is very high, while the coverage in less densely built areas varies. This, to a large degree, reflects issues with accessibility, for example to forested areas, swamps and fields. However, it may also result from fieldworkers’ deprioritization of water bodies located outside the town. In future interventions we recommend testing the accessibility to these areas and emphasizing the importance of identifying water bodies in agricultural areas surrounding neighborhoods.

Overall, fieldworkers detected 4,063 water bodies during the operation, which were treated a total of 23,927 times; in other words, each water body was treated 5.88 times on average. The number of water bodies detected represents a remarkable increase compared to a previous LSM operation in the same area conducted by AGAMal a year earlier without the use of the app, where only a few hundred water bodies were detected. The substantial enhancement in coverage can be ascribed to two main factors. Firstly, the app’s ability to provide extensive service within the individual neighborhoods. Secondly, the crucial role of the dashboard in guaranteeing a thorough coverage of all neighborhoods. Importantly, during the operation, the dashboard identified an overlooked neighborhood, which was then subsequently treated appropriately.

### Effect on malaria cases and mosquito population

During the intervention period, traps in the control area showed an average increase of 65% in the number of mosquitoes caught (median ratio of 1.65), while traps in the intervention area showed a decrease of 38% (median ratio 0.62), which reflects an effect of 62% (ratio between medians of 0.38, 95% CI: (0.17 - 0.63)). Despite the reduction in the mosquito population, no decrease in malaria cases was recorded.

In the intervention arm we excluded from the analysis 4 collection sites where the overall mosquito count in both periods (both before the intervention and after the intervention) was fewer than 5. The counts in these sites were 0 (before) and 0 (after) in one site, 2 (before) and 3 (after) in another, and 4 (before) and 0 (after) in two additional sites. There were no such sites in the control arm, thus all control sites were included in the analysis. The results are not sensitive to the usage of average or median (ratio between averages is 0.44), nor to the exclusion of sites where fewer than 5 mosquitoes were caught over the 15 collection nights in both periods (before and after the intervention; ratio between medians without exclusion is 0.18, ratio of averages 0.39).

There are some limitations to the results. Most notably, while the intervention was carried out in an urban and peri-urban area, the control was in a more rural setting. Additionally, the *Anopheles funestus* species was dominant in the intervention area, constituting about 60% of the *Anopheles* population, while it was almost non-existent in the control area. Breaking down the impact by the different species reveals a significant reduction of 86% in *Anopheles gambiae* in the intervention area, compared to an increase of more than twofold in the control area. We also saw a 30% reduction in the *Culex* population in the intervention area compared to the control, however, this effect is marginally significant due to the low number of *Culex* in the control area. Surprisingly, we observed over a three-fold increase in the *Anopheles funestus* population within the intervention area. However, this surge is difficult to compare with the control area due to its low number of *funestus* mosquitoes. Total collections are summarized in table 1, and the results for different species and genera are summarized in Table 2.

**Table 1:** Total number of mosquitoes collected in light traps.

| Taxon | Total specimens collected |  |
| --- | --- | --- |
|  | Intervention arm | Control arm |
| *Anopheles* spp. | 1065 | 753 |
| *Anopheles gambiae* | 432 | 674 |
| *Anopheles funestus* | 633 | 79 |
| *Culex* spp. | 2436 | 62 |

**Table 2:** Reduction in mosquito density by taxa.

| Taxon | Median ratio |  | Ratio of median ratios |  |
| --- | --- | --- | --- | --- |
|  | Intervention arm | Control arm | Point estimate | 95% CI |
| *Anopheles* spp. | 0.62 | 1.65 | 0.38 * | (0.17-0.63) |
| *Anopheles gambiae* | 0.14 | 2.15 | 0.06 * | (0.05-0.08) |
| *Anopheles funestus* | 3.2 | 1.29 | 2.49 | (0.67-3.71) |
| *Culex* spp. | 0.49 | 0.7 | 0.7 * | (0.50-0.96) |

\* Statistically significant at 5% level

### Cost

The total cost of the intervention was US $48,961, of which US $18,220 consisted of labor, US $23,686 of equipment and consumables, and US $7,055 of software (see appendix 1). This estimation does not include costs associated with the entomological monitoring of adult mosquitoes through light trap collections. This monitoring technique was solely utilized for impact evaluation and, during the operational period, was not made available to managers for the purpose of enhancing performance.

However, costs associated with larval sampling are included, since it is an integral part of quality assurance in most larviciding operations. While Zzapp’s services – including licenses, training and ongoing support – were provided to the project at no cost, software costs were included in the calculation to provide an idea of the potential cost for future programs. The cost of campaign setup, app licenses and technical support was determined based on the price offered by Zzapp to comparable projects subsequent to this project. The total cost attributable to the mapping phase alone is US $7,529, and to the treatment phase is US $33,019 (excluding costs shared by both phases such as mobile devices and software costs). This cost is much lower than that of IRS in the same area, estimated to be around 1 million USD per round or approximately US $5 per person protected.^[15]^ Based on an estimated population of 208,000 in Obuasi and the surrounding intervention communities, the cost of the larviciding intervention was US $0.24 per person protected.

## Discussion

While a notable decrease in the mosquito population was observed, the reported incidence of malaria did not concurrently diminish. Potential reasons for this could lie within the treatment frequency, which was set at every 14 days. Recent research indicates that treatments every five days could yield significantly better outcomes.^[16]^ Adopting this strategy would have potentially enhanced results substantially, while increasing the total operational cost by less than 50%. Another consideration might be the broader trend observed during 2019/2020, when malaria cases in Obuasi notably increased, which might have masked the effects of the temporary mosquito population reduction. In this context, it is noteworthy that a decrease of 40% in malaria cases was observed in the intervention area compared to the control a month after the treatment ceased.

Regardless, there is considerable scope for improving the effectiveness of digital LSM operations. In this study, we noted a surge in the *funestus* population despite an overall decline in the *Anopheles* population, likely due to their predilection for breeding in marshes on the outskirts of the city^[17]^, which may have been missed by field workers. Consequently, future operations should prioritize the speciation of *Anopheles* larvae collected from breeding sites to better understand the specific breeding grounds of various *Anopheles* species in intervention areas, thereby facilitating more targeted and effective interventions.

Digital system adoption not only bolsters operational efficacy but also enables real-time outcome tracking, laying the groundwork for immediate adjustments and enhancements. By amalgamating the benefits of operational flexibility and efficient management, digitization yields highly cost-effective results. In this operation, the cost per person protected represents a 96% cost-saving compared to IRS in the same area, managed by AGAMal. Given its low cost, especially in urban settings, digitally managed LSM could be incorporated into IRS operations with minimal impact on overall costs, providing a comprehensive solution for mosquito control.

Moreover, by capitalizing on the digitization of LSM operations, a comprehensive strategy for mosquito control becomes feasible. Knowledge of the exact locations of water bodies – one of the valuable outputs of digitization – is instrumental in formulating various other vector control activities, such as positioning mosquito traps or earmarking houses for IRS based on proximity to the reported mosquito breeding sites. Furthermore, digitization enables real-time environmental monitoring, such as the emergence of new water bodies or fluctuations in mosquito populations. Digitization not only facilitates these diverse activities individually, but also their integration, thereby fostering an IVM approach that can have a significant impact on malaria reduction.

In addition to the operational benefits, digitization also serves as a catalyst for scientific insights. For example, data garnered from this operation was used by Dia et al. to train a model that used topography to predict the amount of water bodies per hectare.^[18]^ This model, leveraging easily available high-resolution Digital Elevation Model (DEM) data, not only streamlines the identification process but also maximizes the efficient use of resources in areas with limited access. This innovative digital approach significantly outperforms earlier models that used topographic variables or supplementary satellite imagery data.

The benefits of digitization are not limited to the operational and scientific domains. The transparency and accountability that digitization brings have implications for public trust and stakeholder buy-in. “Public trust in and engagement with governments are enhanced when data are made available and processes are transparent.^[19]^ This trust and engagement, in turn, are known to increase the success of anti-malaria operations.^[20]^ Likewise, for governments and other stakeholders, especially funding bodies, digitization provides a clear and auditable trail of activities and results, reinforcing accountability and trust in the efficacy of operations.^[21]^

Finally, in terms of cost-effectiveness, digitization can provide invaluable data that allows for a more nuanced understanding of the financial implications of various strategies. The enhanced ability to track and monitor the costs and outcomes of operations allows for a precise calculation of cost-effectiveness, which is of paramount importance given the limited budgets for malaria control operations. Through close monitoring and evaluation, implementers can identify where resources can be better utilized, where efficiencies can be improved, and where cost savings can be made. This can lead to an increase in the scope and scale of future operations, enabling a greater impact on malaria reduction.

## Data Availability

All data produced in the present study are available upon reasonable request to the authors

## Appendix 1

Operation Costs

**Table 1:** Labor Cost.

| Phase | Element | Number of employees | Days | Daily rate (\$) | Subtotal |
| --- | --- | --- | --- | --- | --- |
| Mapping Phase | fieldworkers | 24 | 17 | 6.9 | 2,815 |
|  | Team leaders | 6 | 17 | 6.9 | 704 |
|  | Drivers | 2 | 17 | 5.19 | 176 |
|  | Entomology staff | 1 | 17 | 7.7 | 131** |
|  | AGAMal management time* | 2 |  |  | 1283** |
| Treatment Phase | Fieldworkers | 12 | 69 | 6.9 | 5713 |
|  | Team leaders | 4 | 69 | 7.7 | 2125 |
|  | Drivers | 2 | 69 | 5.19 | 716 |
|  | AGAMal management time* | 2 |  |  | 4026** |
|  | Entomology staff | 1 | 69 | 7.7 | 531** |
| **Total** |  |  |  |  | **18,220** |

\* Division of management time between the mapping and treatment phases is unclear, and was based on the relative duration of each.

**Table 2:** Equipment, Consumables and Software Costs.

| Phase | Element | Units | Capital cost (\$) | Amortization, monthly (5) | Cost per unit (\$) | Total (\$) |
| --- | --- | --- | --- | --- | --- | --- |
| Mapping Phase | Vehicles* | 3 cars x 1 month | 44,000 | 1.66 | 730.4 | 2,191 |
|  | Fuel | 13.5L x 17 days (estimated) | - | - | 1 | 229 |
| Treatment Phase | Vehicles* | 3 cars x 3.5 months | 44,000 | 1.66 | 730.4 | 7,392 |
|  | Fuel | 13.5L x 69 days (estimated) | - | - | 1 | 931 |
|  | Bti: product + air freight** | 6 | - | - | 1241 | 7,392 |
|  | Bti import: port charges | 6 | - | - | 491 | 2,946 |
|  | Pumps and nozzles | 12*6 months | 139 | 1.66 | 2.3 | 167 |
|  | Nose masks | 48 | 1.72 | - | - | 82 |
|  | Overalls | 12 | 25.8 | - | - | 310 |
|  | Hand gloves | 40 packs | 17.2 | - | - | 688 |
| Both Phases | Mobile devices | 26 * 12 months | 120.8 | 2.48*** | 3.8 | 1,048 |
|  | Boots | 12 | 25.8 | - | - | 310 |
| Software costs | Zzapp: annual app licenses | 15 | - | - | 37 | 555 |
|  | Zzapp: monthly app licenses for mapping phase | 15 | - | - | 100 | 1,500 |
|  | Zzapp: campaign setup, training and technical support | Fee for a small campaign with training on location | - | - | 5,000 | 5,000 |
| **Total** |  |  |  |  |  | **30,741** |

\* Vehicles used for the operation were Toyota Land Cruiser (2010-2014) troop carriers owned by AGAMal. The costing was based on the purchase price of the vehicles, rather than rental prices, which would result in a higher estimate. This is reasonable in the context of an existing malaria control program that chooses to introduce larviciding as an off-season intervention, or a long-term intervention able to purchase the vehicles. The costing was based on 5-year amortization.
\*\* Due to limited supply of VectoBac® WG in Ghana at the time of the operation, the product was air freighted from the manufacturer in the USA. When available locally or if freighted by sea, the cost of the product is significantly lower. The quantity purchased was more than actually used, and the specified amortized cost represents only the portion of the product that was used.
\*\*\* Assuming the devices last through three years of use.

## Cost sensitivity

Possible ways of enhancing the impact of the intervention include increasing its duration, improving the coverage of water bodies in the mapping phase, increasing the water body treatment frequency (when appropriate), and locating more of the water bodies that form after the initial mapping phase. The impact of these changes on cost can be calculated from the tables above.

For example, extending the treatment phase by 1.5 months, without remapping, will increase the cost of the intervention by $13,207 (40% of the cost of the treatment phase). Increasing the frequency of treatment from 14 days to 10 days will have a similar effect on cost. Remapping the area to locate newly formed water bodies will double the cost of mapping, adding $7,529 to the general operation cost.

